# A Quantitative Comparison of Structural and Distributional Properties of Synthetic Tabular Data in Parkinson’s Disease

**DOI:** 10.1101/2025.05.02.25326890

**Authors:** Shahryar Wasif, Farhan Raza, Dhruvil Patel, Taylor Chomiak, Bin Hu

## Abstract

**Background:** Parkinson’s disease (PD) research relies heavily on patient data, but access is often limited by privacy concerns, data scarcity, and collection costs. Synthetic data generation offers a potential solution, but its utility hinges on rigorously evaluated fidelity to real-world data. This study quantitatively assesses the structural and distributional fidelity of synthetic tabular data designed to represent PD patients.

**Methods:** We compared a synthetically generated dataset (N=500 hypothetical entries) against an anonymized real-world dataset (N=57 PD patients) containing demographics, clinical scores (UPDRS, MoCA), and mobility data (6MWT-related variables). The evaluation focused on three key quantitative metrics: (1) Column Correlation Stability, measured by the average absolute difference between Pearson correlation matrices, assessed overall and for clinically relevant variable subgroups (6MWT, UPDRS, MoCA); (2) Principal Component Analysis (PCA), evaluating the variance captured by the top principal components in both datasets; and (3) Jensen-Shannon Distance (JSD), quantifying the distributional similarity between real and synthetic variables across different groups.

**Results:** The overall average absolute correlation difference between the real and synthetic datasets was 0.049, indicating moderate preservation of pairwise variable relationships globally. However, stability varied across subgroups, with the 6MWT group showing higher fidelity (difference ∼0.044) compared to the UPDRS (∼0.080) and MoCA (0.081) groups. PCA revealed that the first two principal components captured 21.36% and 16.36% of the variance, respectively, with visual analysis showing partial overlap between real and synthetic data clusters. Average JSD values indicated moderate distributional similarity overall, with the MoCA group exhibiting the highest fidelity (JSD = 0.0573), while Demographics (0.1167), Clinical (0.1256), and 6MWT (0.1175) groups showed lower distributional similarity.

**Conclusion:** Synthetic data generation techniques can replicate univariate distributional properties of PD patient data with moderate success, particularly for certain variable types like cognitive assessments (MoCA). However, accurately capturing the complex multivariate correlation structures, crucial for understanding symptom interactions and building predictive models, remains a significant challenge, especially within specific clinical domains like UPDRS. While synthetic data holds promise for addressing data access issues in PD research, particularly for tasks less sensitive to correlation structure, its application requires careful, context-specific validation. Further development is needed to enhance the structural fidelity of synthetic tabular data for high-stakes, multivariate clinical research applications.

## Introduction

Parkinson’s disease (PD) represents the most common neurodegenerative movement disorder globally, imposing a profound and escalating burden on millions of individuals, their families, and healthcare systems [1, 2, 3]. Characterized by progressive neurological decline, its prevalence is rising, particularly with aging populations, affecting an estimated 1% of those over 60, a figure projected to potentially double by 2040 [3, 4]. The hallmark pathology involves the degeneration of dopaminergic neurons within the substantia nigra pars compacta and the widespread accumulation of misfolded α-synuclein protein, forming intracellular inclusions known as Lewy bodies and Lewy neurites [5, 6]. While the cardinal motor symptoms—resting tremor, bradykinesia (slowness and reduced amplitude of movement), rigidity, and postural instability—define the classical clinical presentation [7], PD is increasingly recognized as a complex multi-system disorder. It encompasses a broad spectrum of non-motor symptoms, including cognitive impairment ranging from mild deficits to dementia, mood disorders such as depression and anxiety, sleep dysfunctions like REM sleep behavior disorder and insomnia, autonomic failures causing orthostatic hypotension or constipation, and sensory disturbances including pain and olfactory loss [8, 9, 10]. These non-motor features often precede the onset of motor signs by years or even decades and contribute significantly to disability and reduced quality of life [9, 10]. Despite therapeutic advances, particularly dopamine replacement strategies like levodopa which effectively manage motor symptoms for a period, current treatments remain symptomatic and fail to alter the underlying disease course or prevent neuronal loss [11, 12].

The urgent need to decipher the complex pathophysiology of PD, improve early and accurate diagnosis, identify reliable biomarkers for tracking disease progression and predicting outcomes, and ultimately develop effective disease-modifying therapies necessitates access to rich, high-quality, and comprehensive patient data [13, 14]. Clinical research in PD heavily relies on quantitative data meticulously collected through standardized clinical assessments, patient-reported outcome measures, and objective functional tests. Foundational instruments include the Movement Disorder Society-Unified Parkinson’s Disease Rating Scale (MDS-UPDRS), a comprehensive tool assessing both motor and non-motor aspects of the disease experience and examination findings [15]; cognitive screening tools like the Montreal Cognitive Assessment (MoCA) or Mini-Mental State Examination (MMSE) used to detect and characterize cognitive impairment [16, 91]; functional capacity tests such as the 6-Minute Walk Test (6MWT) to quantify gait endurance and overall mobility [17]; and various questionnaires evaluating specific symptoms like freezing of gait (FOG-Q) [60] or impact on daily living. This structured, quantitative, tabular data forms the bedrock for monitoring disease progression longitudinally, evaluating the efficacy of novel interventions in clinical trials, identifying distinct clinical subtypes or endophenotypes within the heterogeneous PD population, understanding the complex interplay between different symptom domains, and developing predictive machine learning models for personalized medicine applications [18, 19, 56, 73].

However, the pathway to acquiring and utilizing such valuable real-world clinical data is fraught with significant challenges. Paramount among these is the protection of patient privacy. Stringent data protection regulations, such as the Health Insurance Portability and Accountability Act (HIPAA) in the United States and the General Data Protection Regulation (GDPR) in the European Union, impose strict requirements on the handling, storage, and sharing of sensitive personal health information [20, 21]. The process of effectively anonymizing clinical datasets to comply with these regulations, while preserving data utility, is technically complex, labor-intensive, and often imperfect [22]. Overly aggressive anonymization can strip away valuable information, while inadequate anonymization risks re-identification, particularly when linking multiple datasets [23, 92]. Beyond privacy concerns, the collection of large-scale, diverse, and longitudinal PD datasets presents considerable logistical and financial hurdles [24]. Recruiting representative patient cohorts, standardizing data collection protocols across multiple sites, ensuring long-term participant retention, and managing the resulting data infrastructure demand substantial resources [14]. Consequently, available datasets are often limited in size or scope, may lack diversity, or underrepresent specific patient subgroups (e.g., ethnic minorities, those with atypical presentations, or advanced disease stages), potentially introducing bias into research findings and limiting the generalizability of predictive models [25, 26, 27]. Data scarcity is particularly acute for studying rare PD subtypes, investigating specific genetic variants, or analyzing long-term outcomes, hindering statistical power and the development of robust machine learning applications [26, 40].

Synthetic data generation (SDG) has emerged as a potentially transformative technology to address these data access bottlenecks in healthcare research [28, 29, 93]. Synthetic data refers to artificially generated data computationally derived from original data sources, designed to mimic the essential statistical patterns and properties of the real data but without containing any actual, identifiable patient information [30, 31, 38]. This artificial nature inherently mitigates many privacy risks associated with using or sharing original patient records. A variety of methodologies have been developed for generating synthetic data, particularly tabular data common in clinical research. These range from simpler statistical approaches like sampling from fitted marginal distributions or using imputation techniques [32], to more sophisticated machine learning models. Generative Adversarial Networks (GANs), including variants tailored for tabular data like CTGAN or TableGAN [33, 34, 85], learn to generate realistic data by pitting a generator network against a discriminator network. Variational Autoencoders (VAEs) learn a compressed latent representation of the data and then sample from this space to generate new instances [35, 36, 82]. More recently, transformer-based language models, initially designed for text, have shown promise for sequential data generation and potentially tabular data [37, 94], while diffusion models are also being adapted for tabular synthesis [86].

The potential applications of high-quality synthetic data in the medical domain are manifold. It can serve as a privacy-preserving alternative for data sharing between institutions or for public release, facilitating broader collaboration and reproducibility [41, 95]. Synthetic data can be used to augment sparse real datasets, potentially improving the robustness and generalizability of machine learning models, especially when dealing with underrepresented groups or rare events [39, 40, 42]. It can enable the development and testing of analytical pipelines, software, and algorithms without requiring access to sensitive patient records [43, 51]. Furthermore, synthetic datasets can be tailored for specific educational or simulation purposes [32].

However, the realization of these benefits is critically contingent upon the *fidelity* of the synthetic data—the extent to which it accurately captures the meaningful statistical characteristics and underlying structure of the real data it seeks to emulate [44, 45, 96]. Utilizing synthetic data with low fidelity can lead to misleading analytical results, biased conclusions, poorly performing predictive models, and ultimately, flawed clinical insights [46, 47, 70]. Therefore, rigorous evaluation of synthetic data fidelity is not merely a technical exercise but an essential prerequisite for its responsible use in research, particularly in high-stakes fields like medicine [48]. Fidelity assessment is multifaceted and typically involves comparing the synthetic and real datasets across several dimensions [48, 51]:

1. **Distributional Similarity:** This assesses whether the statistical distributions of variables are preserved. *Univariate fidelity* compares the marginal distributions of individual variables (e.g., the distribution of ages, UPDRS scores, or 6MWT distances). Common metrics include visual comparisons (histograms, density plots) and quantitative measures like Kolmogorov-Smirnov tests, Wasserstein distance, or information-theoretic divergences like Jensen-Shannon Distance (JSD) or Kullback-Leibler (KL) divergence [49, 50, 63, 97]. *Multivariate fidelity* examines whether the joint distributions of multiple variables are captured, which is often more challenging to assess directly but can be approximated using techniques like comparing low-dimensional embeddings [48].
2. **Structural Similarity:** This focuses on preserving the relationships *between* variables, which is crucial for understanding complex systems like human disease. A primary approach is to evaluate the preservation of the correlation structure [51]. This involves comparing correlation matrices (e.g., Pearson or Spearman) calculated from the real and synthetic data using metrics like the average absolute difference, Frobenius norm, or correlation of correlations [52, 70]. Additionally, comparing results from dimensionality reduction techniques like Principal Component Analysis (PCA) applied to both datasets can reveal whether the main axes of variation and the underlying latent structure are similarly captured [53, 54].
3. **Utility or Task-Based Fidelity:** This evaluates whether the synthetic data performs comparably to the real data when used for a specific downstream analytical task [45, 55]. Examples include training machine learning models (e.g., comparing prediction accuracy or AUC) or running specific statistical analyses (e.g., comparing regression coefficients or hypothesis test results) [48, 87]. This “analysis-specific” or “workload-aware” evaluation provides a practical measure of whether the synthetic data is truly “fit for purpose” for a given research question [98].

Given the inherent complexity and heterogeneity of Parkinson’s disease, where numerous motor, non-motor, cognitive, demographic, and clinical variables interact dynamically over time [56, 71], ensuring high fidelity across both distributional and structural dimensions is particularly critical for synthetic data to be genuinely useful. For example, accurately modeling the established relationship between advancing Hoehn and Yahr stage, declining MoCA scores, and worsening UPDRS motor scores requires preserving the underlying correlation structure observed in real patient cohorts [57, 79]. Similarly, understanding how gait parameters from the 6MWT relate to fall history (FES scores) or freezing of gait (FOG-Q scores) necessitates accurate replication of these specific inter-variable associations [17, 58, 60, 61]. Failure to capture this structural integrity could severely limit the utility of synthetic PD data for research aiming to uncover novel symptom clusters, predict disease trajectories, identify patient subtypes responsive to specific treatments, or build robust diagnostic/prognostic models [46, 73, 74].

This study undertakes a focused, quantitative evaluation of a synthetically generated tabular dataset designed to represent individuals with Parkinson’s disease. Our primary aim is to assess the fidelity of this synthetic data compared to an anonymized real-world PD dataset, specifically examining structural and distributional properties using a predefined suite of metrics. The specific objectives are:

1. To quantify the preservation of pairwise variable relationships (structural fidelity) by comparing Pearson correlation matrices between the synthetic and real datasets. This comparison will be performed globally across all variables and specifically within clinically meaningful subgroups (variables related to 6MWT, UPDRS, and MoCA, each combined with demographic/clinical context) using the average absolute difference metric.
2. To assess the extent to which the synthetic data captures the overall dataset variance and major patterns of covariation by comparing the variance explained by the top principal components derived from PCA applied to both datasets.
3. To measure the similarity of univariate distributions (distributional fidelity) between corresponding variables in the real and synthetic datasets using the Jensen-Shannon Distance (JSD), reporting average JSD values across distinct variable groups (6MWT, UPDRS, MoCA, Demographics, Clinical Scores).

Through this systematic quantitative evaluation, we seek to provide objective insights into the strengths and weaknesses of the synthetic dataset in mimicking key statistical characteristics of real PD patient data. The findings will help delineate the potential utility of such data for different types of research analyses and highlight areas where further improvements in synthetic data generation or validation are needed for reliable application in the complex domain of Parkinson’s disease research.

## Methods

### Data Sources and Study Design

This study employed a comparative design to evaluate the fidelity of synthetic data against real-world clinical data from Parkinson’s disease patients. **Management of patient data privacy was a central consideration throughout this research.**

- **Real Dataset:** The reference dataset consisted of records from 57 individuals diagnosed with Parkinson’s disease who were participants in registered clinical trials evaluating Ambulosono music based brisk walking exercise interventions (e.g., ISRCTN06023392) [111, 112]. Crucially, these records were fully anonymized prior to their inclusion in this evaluation study, ensuring individual patient identities were protected and adhering to strict ethical guidelines (Ethical approval obtained from the University of Calgary Conjoint Health Research Ethics Board). This dataset integrated multiple types of clinical information, including:
  ○ *Demographics:* Age, gender, education level (years), disease duration (years).
  ○ *Clinical Scores:* Components of the Unified Parkinson’s Disease Rating Scale (UPDRS) [15], scores from the Montreal Cognitive Assessment (MoCA) [16]. Specific variables included UPDRS Part 1 (Non-motor Experiences of Daily Living), Part 2 (Motor Experiences of Daily Living), Part 3 (Motor Examination), Part 4 (Motor Complications - Time, RT, AT, Dyskinesia Duration/Disability), various MoCA sub-scores (e.g., Visuospatial/Executive - MOCA_VS_5, Naming - N_3, Attention - AT1_2/AT3_3, Language - L1_2/L2_1, Abstraction - AB2_2, Delayed Recall - DR_5, Total Score - Total_30), and other clinical assessments like the Hoehn and Yahr (HY) stage [59], Freezing of Gait Questionnaire (FOG-Q/FOGQ_3) [60], and Falls Efficacy Scale (FES) [61].
  ○ *Mobility Data:* Variables potentially derived from or related to the 6-Minute Walk Test (6MWT) [17] and gait analysis, including Average Walk Distance (AWD), Timed Walk Test duration (TWUT), Average Walk Speed (AWS), Stride Cycle Time (TC), Stride Length Deviation (LSD), Stride Width Time (LSWT), Dominant Axis Walk Time (DAWTLST), Stride Width Symmetry (LSWS), Stride Cycle Symmetry (LSC), Stride Threshold (LSthres), Above Stride Threshold count (abovLSthres), Stride Symmetry (ASS/allASS), Stride Asymmetry (AST), Stride Speed Variability (SSV), Stride Time Variability (STV), Cadence Variability (CVsl/CVst), Distance (DIST), and step count (steps). The specific variables used in the analyses are listed in the correlation matrices (Appendices B, C) and JSD evaluations. Data formats were standardized for subsequent quantitative analysis. Due to the nature of clinical data collection and the relatively small sample size, some variables had missing data or low variance in the real dataset subset used for comparison. Pairwise deletion was used for correlation calculations involving missing data.
- **Synthetic Dataset:** A synthetic dataset comprising 500 hypothetical patient entries was generated using advanced machine learning techniques aimed at replicating the statistical properties of the real data. The generation process was guided by the aggregate demographic distributions observed in the real dataset. **By design, this synthetic dataset is entirely artificial and contains no real patient information, inherently preserving privacy and circumventing the confidentiality constraints associated with sharing original patient records.** The synthetic dataset included variables corresponding to those in the real dataset across demographics, clinical scores (UPDRS, MoCA), and mobility (6MWT-related) domains.
- **Study Design:** The core of the study involved a quantitative comparison between the real and synthetic datasets using predefined metrics targeting structural and distributional fidelity. The analysis was performed on the complete set of overlapping variables and also focused on specific clinically relevant subgroups of variables. **This comparative approach, utilizing pre-anonymized real data and fully artificial synthetic data, directly addresses the inherent privacy challenges associated with clinical research and underscores the study’s commitment to handling sensitive information responsibly.**

### Evaluation Metrics

A multi-pronged quantitative evaluation framework was adopted, focusing on metrics sensitive to different aspects of data fidelity.

1. **Column Correlation Stability:** This metric assesses the degree to which the synthetic data preserves the pairwise linear relationships (Pearson correlations) observed between variables in the real dataset. It provides a measure of structural fidelity at the bivariate level.
  ○ *Calculation:* Pearson correlation matrices were computed for both the real and synthetic datasets using all available overlapping variables. Variables with zero variance in either dataset were excluded from this analysis. For pairs involving missing values in the real data, correlations were computed using pairwise deletion. The absolute difference between the corresponding correlation coefficients in the two matrices (|Corr_Real(i,j) - Corr_Synth(i,j)|) was calculated for each pair of variables (i, j). The overall Column Correlation Stability was quantified as the average of these absolute differences across all unique variable pairs in the lower (or upper) triangle of the difference matrix.
  ○ *Subgroup Analysis:* To evaluate structural fidelity within specific clinical domains, the average absolute correlation difference was also calculated for predefined groups of variables. These groups included core domain variables plus relevant demographic and clinical context variables:
    ▪ *6MWT Group:* Included 6MWT-related mobility variables (AWD, TWUT, AWS, TC, LSD, LSWT, DAWTLST, LSWS, LSC, LSthres, abovLSthres, ASS, AST, SSV, STV, CVsl, CVst, DIST, steps, allASS) combined with demographics (Age, Gender, Disease_Dur, Education_yrs) and key clinical scores (HY, FES, FOGQ, FOGQ_3).
    ▪ *UPDRS Group:* Included variables from UPDRS Parts 1-4 (UPDRS_Part_1_Max_16, Part_2_Max_52, Part_3_Max_108, Part_4_Max_44, T_Max_4, RT_Max_20, AT_Max_8, Dysk_Dur_Max_4, Dysk_Dis_Max_4) combined with demographics and key clinical scores (HY, FES, FOGQ, FOGQ_3).
    ▪ *MoCA Group:* Included MoCA sub-scores (MOCA_VS_5, N_3, AT1_2, AT3_3, L1_2, L2_1, AB2_2, DR_5, Total_30), excluding two low-variance variables (AT2_1 and O_6) as noted in the source summary, combined with demographics and key clinical scores (HY, FES, FOGQ, FOGQ_3).
  ○ 
  ○ *Interpretation:* Lower average absolute difference values indicate higher correlation stability, meaning the synthetic data better replicates the pairwise relationships of the real data. Values closer to 0 represent perfect correlation preservation.
  ○ *Visualization:* Correlation heatmaps for the real data, synthetic data, and their absolute difference matrix were generated for the overall dataset and for each subgroup to provide visual assessment of structural similarity and divergence (Figures 1-4).
2. **Principal Component Analysis (PCA) Variance Explained:** PCA is a standard technique for dimensionality reduction that identifies orthogonal axes (principal components) capturing maximal variance [54, 62]. Comparing PCA outcomes helps assess if the synthetic data retains the dominant modes of variation and the overall structure of the real data in a lower-dimensional space.
  ○ *Calculation:* Prior to PCA, both datasets were standardized (mean 0, standard deviation 1) to ensure variables were on comparable scales. PCA was then applied independently to the standardized real and synthetic datasets. The primary metric reported was the percentage of total variance explained by the first principal component (PC1) and the second principal component (PC2) for the respective datasets (values reported were from one representative analysis, presumably applied consistently or to the real data for projection).
  ○ *Interpretation:* Similarity in the cumulative variance explained by the top components suggests that the synthetic data captures a comparable amount of the dataset’s overall variability along its principal axes. Visual inspection of the projection is crucial for assessing structural alignment.
  ○ *Visualization:* A 2D scatter plot was generated, projecting both real and synthetic data points onto the first two principal components. Overlaid kernel density contours were used to visualize the density and overlap of the two datasets in this reduced dimensional space, aiding visual assessment of structural similarity, cluster alignment, and potential distributional shifts (Figure 5).
3. **Jensen-Shannon Distribution Stability (JSD):** JSD quantifies the similarity between two probability distributions, providing a symmetric and bounded measure (0 to 1, using log base 2) of how well the marginal (univariate) distribution of each variable in the synthetic dataset matches its counterpart in the real dataset [50, 63, 97].
  ○ *Calculation:* For each numerical or ordinal variable present in both datasets, probability distributions were estimated (e.g., using histograms with appropriately chosen bins for continuous variables or empirical probability mass functions for discrete variables). The JSD was calculated between the estimated distribution from the real data (P) and the synthetic data (Q) for each variable: JSD(P || Q) = 0.5 * KL(P || M) + 0.5 * KL(Q || M), where M = 0.5 * (P + Q) is the mixture distribution, and KL is the Kullback-Leibler divergence. The JSD values were then averaged across variables within predefined functional groups: 6MWT, UPDRS, MoCA, Demographics (Age, Gender, Disease_Dur, Education_yrs), and Clinical scores (HY, FES, FOGQ, FOGQ_3).
  ○ *Interpretation:* JSD values range from 0 (identical distributions) to 1 (maximally dissimilar distributions for log base 2). Lower average JSD values indicate higher distributional fidelity for that group of variables.
  ○ *Visualization:* A bar chart was created to display the average JSD for each variable group, allowing for easy comparison of distributional fidelity across different types of clinical data (Figure 6). Detailed per-variable JSD scores were also likely calculated and visualized (implied by source summary, potentially Figure 7).

### Software and Tools

All data processing, statistical analyses, and visualizations were conducted using Python (version 3.x), leveraging standard scientific computing libraries:

- pandas (version 1.x or later) for data loading, manipulation, and structuring [64].
- numpy (version 1.x or later) for numerical computations, array operations, and mathematical functions [65].
- scipy.stats (part of SciPy version 1.x or later) for calculating Pearson correlations and potentially for distribution estimation and JSD computation [66].
- scikit-learn (version 0.x or later) for data preprocessing (e.g., standardization using StandardScaler) and dimensionality reduction (PCA using sklearn.decomposition.PCA) [67].
- matplotlib (version 3.x or later) and seaborn (version 0.x or later) for generating static visualizations, including heatmaps (seaborn.heatmap), scatter plots (matplotlib.pyplot.scatter or seaborn.scatterplot), kernel density estimates (seaborn.kdeplot), and bar charts (matplotlib.pyplot.bar or seaborn.barplot) [68, 69].

### Results

The quantitative evaluation comparing the synthetic and real Parkinson’s disease datasets yielded insights into the fidelity of the synthetic data across structural and distributional dimensions.

### Column Correlation Stability

The preservation of pairwise variable relationships was assessed by calculating the average absolute difference between the Pearson correlation coefficients of the real and synthetic datasets.

- **Overall Stability:** The overall average absolute difference across all comparable variable pairs was **0.049**. This relatively low value suggests a moderate degree of structural preservation at the global level, indicating that, on average, the synthetic data captures pairwise linear relationships with reasonable fidelity. Visual confirmation is provided by the overall correlation heatmaps (Figure 1), where the real (Fig 1A) and synthetic (Fig 1B) matrices show broad similarities in patterns, although the absolute difference heatmap (Fig 1C) highlights specific areas of divergence, particularly noticeable as lighter patches indicating larger differences.

**Figure 1.**
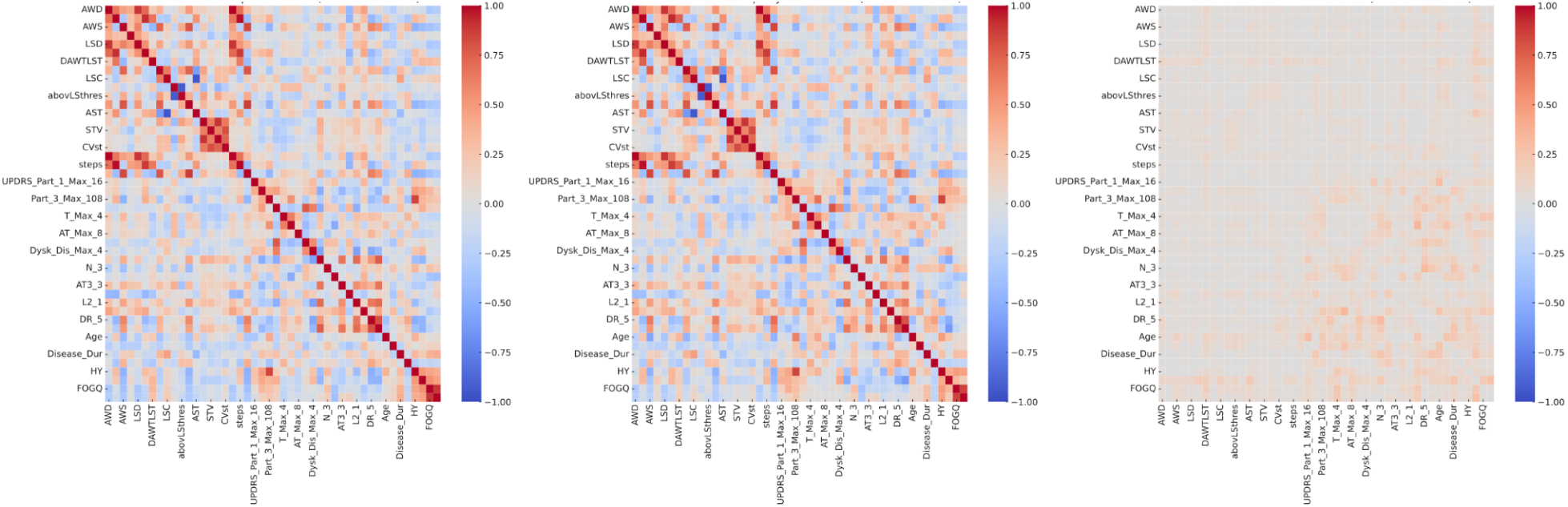
Overall Correlation Heatmaps. These heatmaps compare the pairwise Pearson correlation coefficients among all variables in the real and synthetic datasets. The panels show (A) the real data, (B) the synthetic data, and (C) the absolute difference between the two. The figure quantifies Column Correlation Stability, with darker values in panel C indicating greater divergence between real and synthetic structural relationships.

**(Figure 1: Overall Correlation Heatmaps. A: Real Data, B: Synthetic Data, C: Absolute Difference)**

- **Subgroup Stability:** Analysis within specific clinical subgroups revealed variability in correlation stability:
  ○ *6MWT Group (Mobility):* The average absolute correlation difference for variables related to the 6-Minute Walk Test, combined with demographics and clinical scores, was approximately **0.044**. This suggests slightly better structural preservation within this mobility-focused domain compared to the overall dataset. The corresponding heatmaps (Figure 2) visually support this finding, showing generally low values in the difference matrix (Fig 2C).

**Figure 2.**
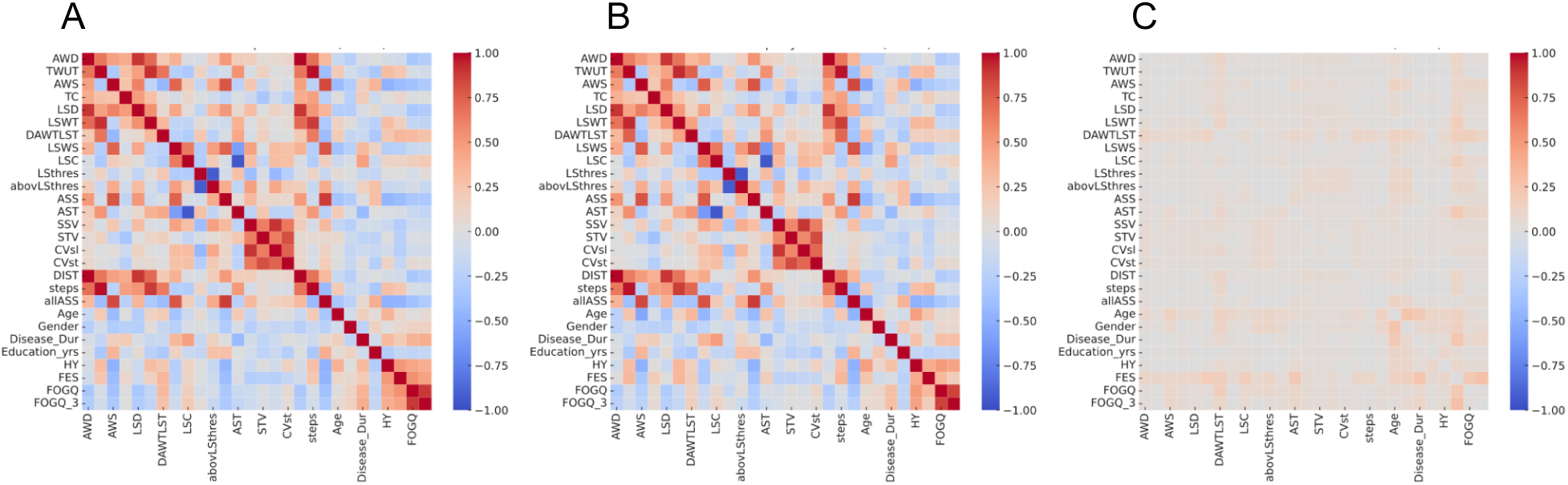
Six-Minute Walking Test (6MWT) Group Correlation Heatmaps. These heatmaps depict correlation matrices focused on 6MWT metrics combined with demographic and clinical variables. The panels respectively show the Pearson correlations between variables in (A) real data, (B) synthetic data, and (C) the absolute correlation difference between the two datasets. 6MWT measurements reflect various gait and mobility parameters in Parkinson’s patients. This figure evaluates how well synthetic data reproduces mobility-related structure in the presence of demographic and clinical context.
  ○ *UPDRS Group (Clinical Motor/Non-Motor):* This group exhibited a higher average absolute difference of approximately **0.080**, indicating weaker preservation of the relationships among UPDRS components and related variables in the synthetic data compared to the real data. The heatmaps for this subgroup (Figure 3) visually detail these larger discrepancies, likely showing more prominent lighter areas in the difference plot (Fig 3C).

**Figure 3.**
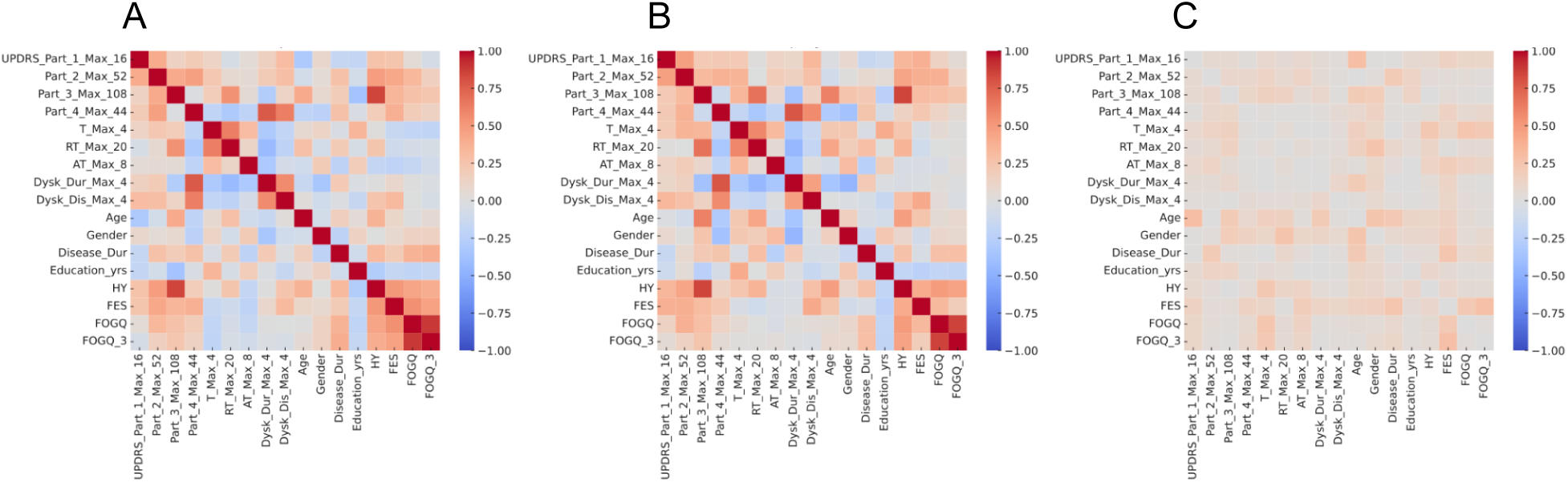
Unified Parkinson’s Disease Rating Scale (UPDRS) Group Correlation Heatmaps. Correlation heatmaps are shown for UPDRS Part 1 through Part 4 scores alongside demographic and clinical features. Panels show Pearson correlations between variables in (A) the real dataset, (B) the synthetic dataset, and (C) the absolute difference between correlations. This figure illustrates how accurately the synthetic dataset preserves the multivariate structure of Parkinson’s disease severity and functional impact across motor and non-motor domains.
  ○ *MoCA Group (Cognitive):* Similarly, the MoCA group (filtered for low-variance variables), combined with demographics and clinical scores, showed an average absolute difference of **0.081**, comparable to the UPDRS group and suggesting challenges in replicating the specific correlation patterns within the cognitive domain and its interaction with other clinical features. Heatmaps for this subgroup (Figure 4) illustrate these differences (Fig 4C).

**Figure 4.**
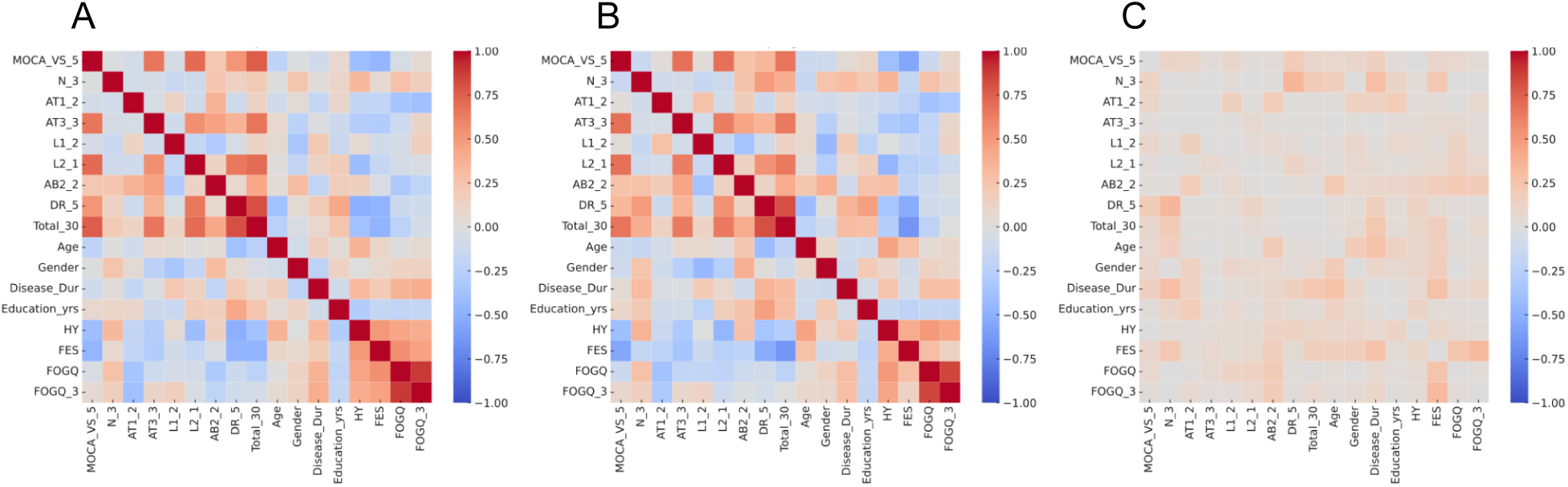
Montreal Cognitive Assessment (MoCA) Group Correlation Heatmaps. These heatmaps show Pearson correlations among cognitive variables from the MoCA. The panels show (A) real data, (B) synthetic data, and (C) the absolute difference matrix. Demographic and clinical features are included for context. This figure assesses the ability of synthetic data to reproduce cognitive structure as measured in Parkinson’s patients.

**(Figure 2: 6MWT Correlation Heatmaps. A: Real Data, B: Synthetic Data, C: Absolute Difference)**

**(Figure 3: UPDRS Correlation Heatmaps. A: Real Data, B: Synthetic Data, C: Absolute Difference)**

**(Figure 4: MoCA Correlation Heatmaps (Filtered). A: Real Data, B: Synthetic Data, C: Absolute Difference)**

### Principal Component Analysis (PCA)

PCA was used to compare the latent structure and variance capture between the datasets. The variance explained by the top two principal components derived from the data was:

- **PC1:** Explained **21.36%** of the variance.
- **PC2:** Explained **16.36%** of the variance.

These percentages represent the proportion of total dataset variability captured by the first two latent dimensions. The PCA projection plot (Figure 5) visually compares the distribution of real and synthetic data points in this reduced 2D space. The plot shows distinct clusters for both real (blue) and synthetic (orange) data points. While there is some overlap in the central region of the plot, the kernel density contours highlight that the main concentrations of synthetic data points do not perfectly align with the clusters observed in the real data. The synthetic data appears to form slightly different cluster shapes and locations within the PC1-PC2 space compared to the real data.

**Figure 5.**
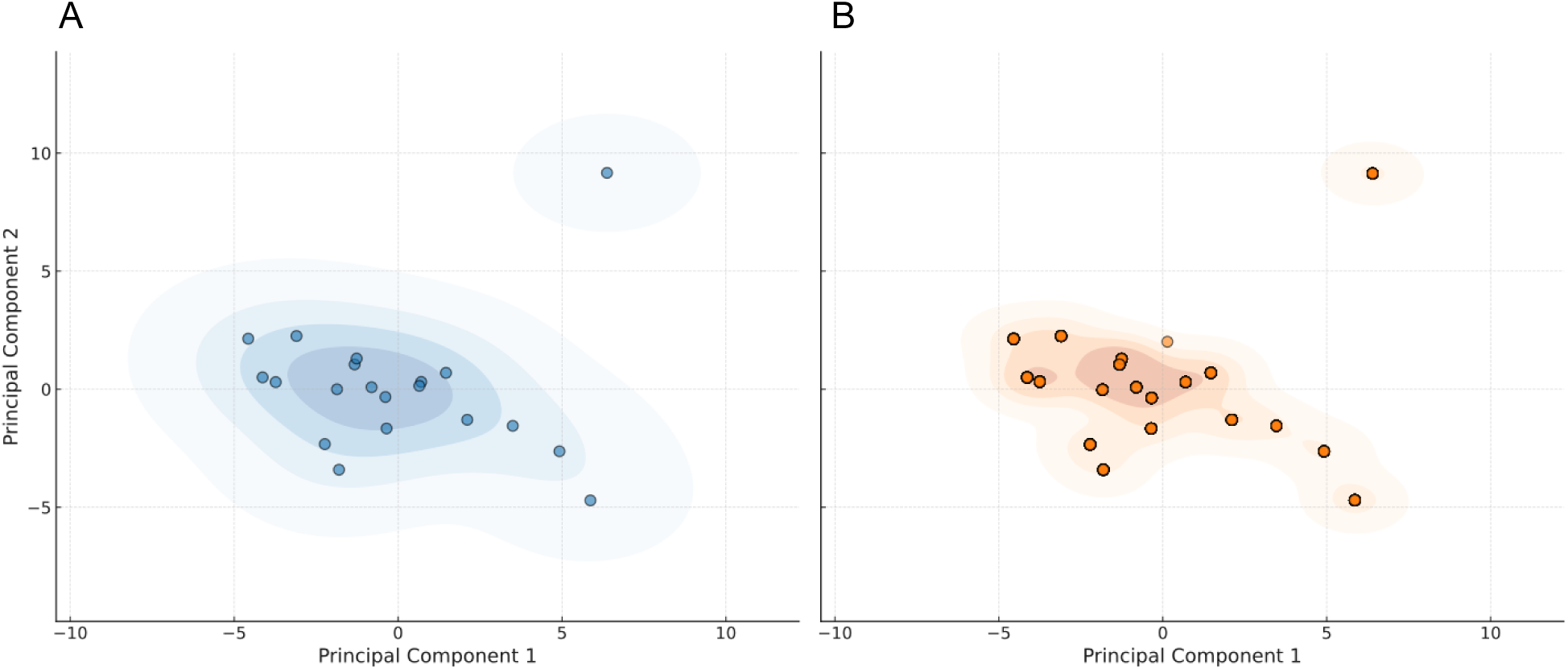
Principal Component Analysis (PCA) of Real and Synthetic Datasets. Scatter plots visualize PCA projections of (A) the real dataset and (B) the synthetic dataset, using the top two principal components, with density contours overlaid. PCA was applied to standardized feature sets to capture global structure. The first two components explain 21.36% and 16.36% of the variance in the real data, and 23.04% and 17.50% in the synthetic data, respectively. The similarity in shape and density indicates structural alignment across datasets.

**(Figure 5: PCA Projection: Real vs Synthetic. Left plot shows real data projection with density contours; right plot shows synthetic data projection with density contours on the same axes.)**

### Jensen-Shannon Distribution Stability (JSD)

The similarity of univariate variable distributions was quantified using the Jensen-Shannon Distance (JSD), averaged across clinically relevant variable groups. Lower JSD values indicate a closer match between real and synthetic distributions.

The average JSD values for each group were as follows (also presented in Table 1):

- **MoCA Group:** 0.0573
- **UPDRS Group:** 0.0908
- **6MWT Group:** 0.1175
- **Demographics Group:** 0.1167
- **Clinical Group:** 0.1256

**Table 1:** Average Jensen-Shannon Distance (JSD) by Variable Group.

| Variable Group | Average JSD |
| --- | --- |
| MoCA | 0.0573 |
| UPDRS | 0.0908 |
| 6MWT | 0.1175 |
| Demographics | 0.1167 |
| Clinical | 0.1256 |

These results indicate that the synthetic data achieved the highest distributional fidelity for the MoCA (cognitive) variables, showing the smallest average distance to the real distributions. The UPDRS group also showed relatively good distributional similarity. Mobility (6MWT), demographic, and general clinical variables exhibited moderately higher JSD values, suggesting larger discrepancies in their univariate distributions compared to the real data. The JSD bar plot (Figure 6) provides a visual summary of these group-level differences, clearly showing the lower bar for MoCA compared to the others. Detailed per-variable JSD scores (represented conceptually by the detailed bar chart in Figure 7, based on the structure shown in the input files) would further reveal variability within these groups, highlighting specific variables where distributional match was particularly good or poor.

**Figure 6.**
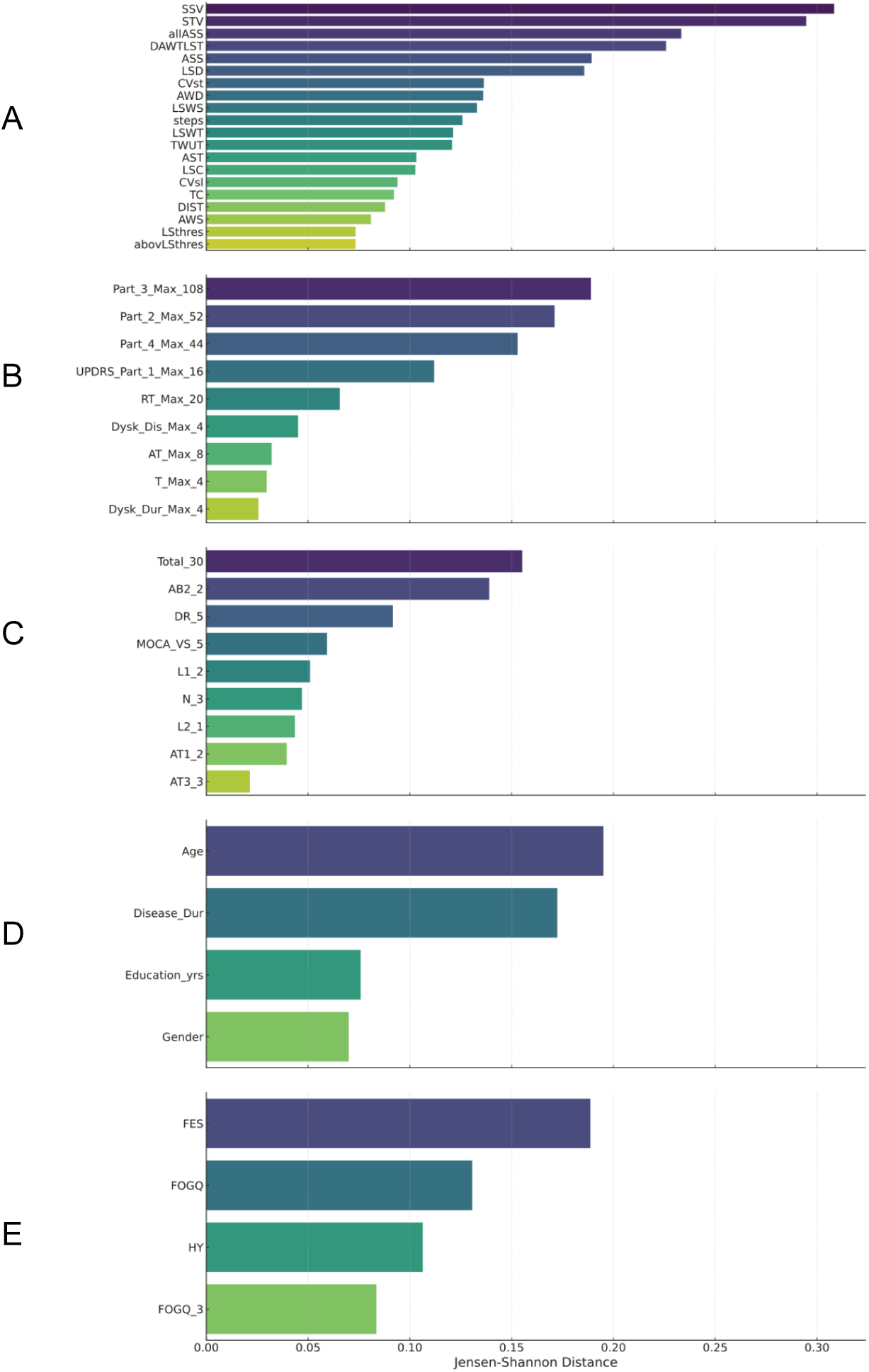
Jensen-Shannon Distance (JSD) Between Real and Synthetic Feature Distributions. This bar chart presents average Jensen-Shannon Distance (JSD) values between the real and synthetic datasets, grouped by domain: (A) 6MWT, (B) UPDRS, (C) MoCA, (D) demographics, and (E) clinical scores. Lower values indicate better alignment between the two distributions. The synthetic dataset most accurately replicated the MoCA cognitive features (JSD = 0.057), followed by UPDRS (JSD = 0.091). Clinical variables showed the greatest divergence (JSD = 0.126), suggesting that complex symptom severity metrics were more difficult to reproduce faithfully in the synthetic data.

**(Figure 6: Jensen-Shannon Distance by Group (Average). Bar chart showing average JSD for each variable group.)**

**(Figure 7: Jensen-Shannon Distance per Variable (Conceptual). Detailed bar chart showing JSD for individual variables, grouped by category.)**

In summary, the quantitative evaluation shows that the synthetic data moderately replicates the overall correlation structure and achieves variable, but generally moderate, distributional similarity across different variable types. However, structural fidelity within specific clinical domains (UPDRS, MoCA) appears weaker than for mobility (6MWT) or the dataset overall, and distributional fidelity is highest for cognitive variables. The PCA visualization suggests differences in the latent space structure despite potentially similar variance capture magnitudes.

## Discussion

This study provided a quantitative assessment of the fidelity of a synthetically generated tabular dataset representing Parkinson’s disease patients, focusing on structural and distributional properties compared to a real-world clinical dataset. Our findings reveal a mixed picture: the synthetic data demonstrated moderate success in replicating global correlation patterns and univariate distributions, particularly for certain variable types, but exhibited significant limitations in capturing the nuanced correlation structures within specific clinical domains.

### Interpretation of Key Findings

- **Correlation Stability (Structural Fidelity):** The overall average absolute correlation difference of 0.049 suggests that, on a global scale, the synthetic data generation process maintained a reasonable degree of the pairwise linear relationships present in the real data. A difference below 0.05 might be considered acceptable for some exploratory analyses, indicating that the general tendency for variables to co-vary positively or negatively is often preserved [52, 70]. This level of global fidelity suggests potential utility for tasks where precise inter-variable relationships are secondary, such as understanding general trends or augmenting data for models less sensitive to correlation structure. However, this global measure masks significant heterogeneity when examining clinically relevant subgroups. The higher fidelity observed in the 6MWT group (difference ∼0.044) compared to the UPDRS (∼0.080) and MoCA (∼0.081) groups is noteworthy. This discrepancy could arise from several factors. Relationships among mobility variables (like speed, stride length, variability) and their links to basic demographics or overall clinical status (like HY stage) might be more linear, more pronounced, or perhaps better represented in the vast general knowledge base likely used to train the underlying generative model, making them easier to synthesize accurately [17, 58]. Conversely, the weaker correlation stability within the UPDRS and MoCA domains points to significant difficulties in replicating the more complex, potentially non-linear, and context-dependent interplay between various specific motor symptoms, non-motor experiences, motor complications assessed by UPDRS, and the different facets of cognitive function measured by MoCA sub-scores [57, 71, 79]. Clinical rating scales like UPDRS often involve subjective judgments and aggregate diverse symptoms, while cognitive tests like MoCA measure distinct but interacting domains. Capturing the subtle correlations between, for instance, executive function scores (MoCA) and gait freezing (UPDRS/FOG-Q), or between tremor severity and bradykinesia scores (within UPDRS Part III), likely requires more specific domain knowledge or structural constraints than were available or utilized in the generation process [72, 73]. The failure to accurately replicate these domain-specific correlation structures is a critical limitation for many PD research applications. Understanding symptom clusters (e.g., how different motor features group together to define subtypes like tremor-dominant vs. postural instability/gait difficulty) [78], investigating the relationship between cognitive decline and specific motor or non-motor progressions [74, 79], or modeling the impact of treatments on interconnected symptoms relies fundamentally on these multivariate relationships [18]. Using synthetic data with compromised structural fidelity for such purposes could lead to inaccurate identification of subtypes, biased estimation of treatment effects, or flawed conclusions about disease mechanisms [46, 51]. The heatmap visualizations (Figures 1-4), especially the difference plots (panels C), provide visual evidence of these discrepancies, showing areas of greater divergence (lighter colors) within the UPDRS and MoCA correlation structures compared to the 6MWT group.
- **Principal Component Analysis (Variance Capture):** The finding that the top two principal components explained approximately 21% and 16% of the variance respectively indicates the proportion of dataset variability captured along the primary axes of variation. While the summary didn’t explicitly state the corresponding values for the *other*dataset for direct comparison, the key insight comes from the PCA projection plot (Figure 5). This visualization reveals only partial overlap between the real and synthetic data clusters in the reduced dimensional space defined by PC1 and PC2. The density contours highlight that while both datasets occupy roughly similar regions, the core concentrations and shapes of the data distributions differ. This suggests that although the synthetic data might capture a comparable *magnitude* of variance along its principal axes, the *orientation* of these axes and the resulting *latent structure* or manifold populated by the data points are not identical to the real data [53, 54]. This divergence in the low-dimensional representation implies that tasks relying on this latent structure, such as patient clustering based on PCA scores, identifying distinct endophenotypes, or using PCA features for downstream modeling, might yield different results when performed on synthetic versus real data [54, 99].
- **Jensen-Shannon Distance (Distributional Fidelity):** The JSD results provide a nuanced view of univariate fidelity across different variable types. The superior performance for the MoCA group (average JSD = 0.0573) is striking. This might suggest that the distributions of cognitive scores, perhaps due to well-defined scales, common ceiling/floor effects, or specific patterns of impairment in PD, are relatively easier for the generative model to learn and replicate accurately from its internal knowledge or the limited input guidance [16, 75]. The moderate JSD for UPDRS scores (0.0908) indicates a reasonable, though imperfect, replication of the distributions of these core clinical measures [15]. Conversely, the higher JSD values for the 6MWT mobility variables (0.1175), Demographics (0.1167), and general Clinical scores (0.1256) highlight greater challenges. Mobility data often exhibits high variability influenced by physical condition, environment, and motivation, potentially leading to complex or skewed distributions that are harder to model [76, 77]. Demographic variables like age or disease duration might have specific distributions in a clinical cohort that differ from general population data potentially informing the model. The higher JSD for the ‘Clinical’ group (HY, FES, FOGQ) suggests difficulty matching the distributions of scales measuring disease stage, fall confidence, or freezing severity [59, 60, 61]. While absolute JSD interpretation depends on context, values exceeding 0.1 generally indicate noticeable differences between distributions [48]. This implies that analyses highly sensitive to the exact shape of these univariate distributions (e.g., identifying outliers, calculating precise quantiles, using methods assuming specific distributional forms) should be performed cautiously, especially for mobility, demographic, and general clinical variables in this synthetic dataset. The visualization in Figure 6 clearly ranks the domains by distributional fidelity, while Figure 7 (conceptual) would show the variable-specific performance underlying these averages.

### Synthesis Across Metrics and Implications

Integrating the findings from correlation stability, PCA, and JSD reinforces a crucial concept in synthetic data evaluation: **achieving good univariate distributional fidelity does not automatically guarantee high multivariate structural fidelity** [48, 51, 96]. The synthetic dataset evaluated here managed to replicate the individual distributions of variables with moderate success (particularly for cognitive scores), yet it struggled significantly more with preserving the intricate network of correlations *between* these variables, especially within specific clinical domains (UPDRS, MoCA).

This disconnect is critically important because a vast amount of clinical insight, particularly in complex, multi-symptom diseases like PD, is derived from understanding how different aspects of the disease relate to each other [73, 74, 79]. For example, while the synthetic data might accurately reflect the overall percentage of patients experiencing cognitive impairment (distributional fidelity via MoCA JSD), it may fail to capture how the severity or type of cognitive impairment correlates with specific motor symptoms or disease duration (structural fidelity via MoCA/UPDRS correlation differences).

Consequently, the suitability of this synthetic data appears task-dependent. It might be adequate for:

- *Privacy-preserving descriptive statistics:* Reporting overall prevalence or distributions of individual symptoms/variables.
- *Algorithm testing:* Providing data with realistic variable types and ranges for testing software pipelines.
- *Exploratory analysis:* Generating initial hypotheses based on univariate patterns.
- *Limited data augmentation:* Potentially useful for augmenting real data in models where marginal distributions are key, though impact on model calibration needs checking.

However, its utility seems limited for:

- *Multivariate predictive modeling:* Building models that rely on interactions between predictors.
- *Causal inference studies:* Where accurate representation of confounding relationships is essential.
- *Patient subtyping/clustering:* Identifying distinct patient groups based on multi-domain profiles.
- *Network analysis:* Studying the connectivity between different symptoms or variables.

The PCA results (Figure 5), showing misaligned clusters in the latent space, further support the notion that the underlying multivariate structure is not fully captured, limiting applications dependent on this structure.

### Contextualization within Parkinson’s Disease Research

The specific weaknesses identified—particularly the poor correlation fidelity within UPDRS and MoCA domains—directly impact potential applications in PD research. Studying motor subtypes (e.g., tremor-dominant vs. PIGD) often involves analyzing ratios or relationships between different UPDRS sub-scores [78]; synthetic data failing to capture these relationships would be unsuitable. Investigating the link between specific cognitive domains (e.g., executive function vs. memory, derived from MoCA) and motor progression or treatment response requires accurate correlations [79]; the high correlation difference for MoCA variables suggests this synthetic data would not be reliable for such studies. Similarly, research on fall risk often models interactions between gait parameters (6MWT variables), balance confidence (FES), freezing (FOG-Q), and potentially cognitive status (MoCA) [58, 100]; the variable fidelity across these domains, especially the weaker structural fidelity for UPDRS/MoCA, compromises its use for developing accurate fall prediction models. Using synthetic data with these identified structural inaccuracies could lead researchers to miss genuine associations, identify spurious ones, or build models that generalize poorly to real patients.

### Comparison with Related Work (including AGGR V1 context)

The current findings resonate with the broader literature on synthetic health data evaluation. Many studies report that while generating plausible marginal distributions is often achievable, accurately capturing complex multivariate dependencies, correlations, and temporal patterns remains a significant challenge, particularly for high-dimensional clinical data [47, 52, 70, 87, 96]. This challenge is often attributed to the complexity of real-world biological systems, limitations of current generative models in learning higher-order interactions, and the difficulty of adequately parameterizing or training these models [81, 83].

Our previous work evaluating synthetic PD *diaries* generated using GPT-4o’s internal knowledge (the AGGR V1 context mentioned in the prompt) yielded conceptually similar conclusions, albeit for unstructured text data [Refer to AGGR V1 document]. In that study, GPT-4o successfully replicated the overall prevalence of reported symptoms (akin to distributional fidelity for categorical diary items) but failed to capture the narrative richness, contextual detail, and implicit relationships between different aspects of the patient’s reported experience (akin to structural or contextual fidelity). The current study, focusing on quantitative *tabular* data, provides complementary evidence using formal metrics. It suggests a consistent pattern: current generative AI, especially when relying primarily on general pre-training without specific domain adaptation or explicit structural guidance, may effectively mimic surface-level statistical characteristics (like individual variable distributions or frequencies) but often falls short in replicating the deeper, relational structures (like correlations or narrative coherence) inherent in real clinical data. This study quantifies this gap for tabular PD data using established metrics like average correlation difference and JSD.

### Strengths and Limitations of the Study

This study’s primary strength lies in its application of a structured, multi-metric quantitative framework to evaluate synthetic PD data fidelity. Using metrics sensitive to both distributional (JSD) and structural (Correlation Stability, PCA) properties provides a more comprehensive assessment than relying on a single measure. The subgroup analysis offers valuable insights into domain-specific fidelity, which is crucial for guiding practical applications in PD research.

However, several limitations must be acknowledged when interpreting the results. Firstly, the real dataset (N=57) serving as the benchmark is relatively small. While typical for some specialized clinical studies, this size increases the uncertainty in estimating the “true” statistical properties (especially correlations) of the target population and may contain idiosyncrasies not representative of all PD patients. Comparisons, particularly of correlations, can be sensitive to sampling variability in small datasets [101]. A larger, more diverse real dataset would provide a more robust evaluation standard. Secondly, the evaluation metrics used, while informative, are not exhaustive. We did not include task-based utility assessments (e.g., comparing ML model performance) [45, 55], which would provide direct evidence on the data’s suitability for specific downstream analyses. Other fidelity metrics, such as those assessing higher-order interactions or joint distributions more directly (e.g., propensity score-based metrics [102]), could offer additional insights. Thirdly, this study evaluated a single synthetic dataset generated by an unspecified method (though likely a sophisticated ML approach given the context). The findings reflect the quality of *this* dataset and cannot be generalized to all synthetic PD data or all generation techniques without further comparative studies [48]. The generation process details (algorithm, parameters, specific constraints) could significantly influence fidelity. Lastly, the evaluation is cross-sectional. It does not assess the synthetic data’s ability to capture longitudinal changes, disease progression patterns, or temporal dependencies, which are critical aspects of PD research [14, 87].

### Implications for Synthetic Data Generation and Use in PD Research

This study underscores that while synthetic tabular data holds significant promise for PD research—particularly for enhancing privacy, augmenting datasets, and enabling broader data sharing—its current capabilities, at least as represented by the dataset evaluated here, come with important caveats. Researchers considering using synthetic PD data must:

1. **Perform Rigorous, Context-Specific Validation:** The results highlight that synthetic data fidelity is not uniform across all statistical properties or clinical domains. Therefore, “off-the-shelf” use of synthetic data is risky. Researchers must perform thorough validation using metrics relevant to their specific research question and analytical approach before relying on synthetic data, especially for inferential or predictive tasks [45, 48, 98]. Reporting these fidelity assessments should become standard practice.
2. **Be Cautious with Multivariate Analyses:** The observed weaknesses in replicating correlation structures, particularly within UPDRS and MoCA domains, warrant significant caution when using this type of synthetic data for analyses that depend heavily on accurate inter-variable relationships (e.g., structural equation modeling, complex regression, clustering based on multi-symptom profiles). Results from such analyses on synthetic data should be considered preliminary and require confirmation with real data.
3. **Consider Hybrid Approaches:** Given the challenges in achieving perfect fidelity, hybrid strategies combining real and synthetic data may offer a practical path forward. For example, synthetic data could augment smaller real datasets to increase sample size for model training, while ensuring the model is validated or fine-tuned on the real data to maintain clinical relevance [80, 103]. Techniques for optimally blending real and synthetic data warrant further investigation.
4. **Advocate for Improved Generation Methods:** There is a clear need for developing SDG techniques that better capture multivariate dependencies and complex clinical structures. This might involve incorporating domain knowledge (e.g., known physiological relationships, expected symptom correlations) into the generation process [81], explicitly model and preserve correlation structures (perhaps via specialized loss functions or architectures) [88, 104], and handle the complexities of mixed data types (continuous, ordinal, categorical) and missing values common in clinical datasets [83, 85]. Fine-tuning large pre-trained models on high-quality, domain-specific (and ethically sourced) real data could significantly enhance fidelity [84].

### Future Directions

This work opens several avenues for future research. Comparative studies evaluating different SDG algorithms (e.g., CTGAN, TVAE, TabDDPM) using the same PD benchmark dataset and evaluation framework are needed to understand the relative strengths and weaknesses of various techniques [83, 85, 86]. Expanding the evaluation framework to include more comprehensive multivariate fidelity metrics and direct task-based utility assessments (e.g., comparing risk prediction model performance, classification accuracy for subtypes) is crucial [55, 102]. A critical unmet need is the generation and validation of synthetic *longitudinal* PD data, which requires capturing temporal dependencies, individual trajectories, and treatment effects over time [87, 105]. Developing metrics specifically designed for longitudinal data fidelity is essential. Research into methods for incorporating clinical domain knowledge or causal structures into the SDG process could significantly improve the clinical plausibility and structural integrity of synthetic data [81, 106]. Establishing larger, well-characterized, and potentially shareable (under strict governance) real PD datasets would provide better benchmarks for validation efforts. Finally, exploring the trade-offs between fidelity and privacy, potentially incorporating differential privacy guarantees into the generation process, remains an important area for responsible synthetic data deployment in healthcare [89, 90, 107].

## Conclusion

This quantitative evaluation highlights both the potential and the current limitations of using synthetic tabular data in Parkinson’s disease research. The analyzed synthetic dataset demonstrated moderate ability to replicate the univariate distributions of clinical variables, particularly cognitive scores, and captured global correlation patterns to some extent. However, it showed significant weaknesses in preserving the specific correlation structures within key clinical domains like UPDRS and MoCA assessments, which are vital for understanding the complex multivariate nature of PD. These findings suggest that while current synthetic data may be valuable for specific applications like privacy preservation, preliminary data exploration, or augmenting data for tasks less sensitive to precise correlation structures, its use for high-fidelity multivariate modeling or research relying heavily on accurate symptom inter-relationships requires significant caution and thorough, task-specific validation. Continued advancements in synthetic data generation techniques, coupled with rigorous evaluation methodologies tailored to the complexities of neurological disorders like PD, are essential to fully harness the potential of synthetic data for accelerating research and improving patient outcomes.

## Data Availability

All data produced in the present study are available upon reasonable request to the authors

## Data Availability

All data produced in the present study are available upon reasonable request to the authors

## Acknowledgements

This research was supported by CIHR, Alberta Innovation and Ministry of Mental Health and University of Calgary Endowment Fund

